# Blood lead concentrations in exposed forecourt attendants and taxi drivers in parts of South Africa

**DOI:** 10.1101/2023.05.14.23289954

**Authors:** J.O. Olowoyo, U.A. Tshoni, A.S. Kobyana, G.N. Lion, L.L. Mugivhisa, L Koski, S.K.T.S. Wärmländer, P.M. Roos

## Abstract

**Background:** Leaded fuel was banned in South Africa in 2006, in order to improve human health and reduce environmental pollution. Lead (Pb) has been suggested to contribute to the development of neurodegenerative disorders, and the role of respiratory exposure to Pb from petrol fumes should not be neglected in this context. In addition to Pb, petrol contains various harmful chemicals including other neurotoxic metals and hydrocarbons.

**Objectives and Methods:** Here, we investigated concentrations of Pb and other metals in blood from taxi drivers (n=21), petrol station forecourt attendants (n=38), and a control group (n=36). Participants were divided into three groups based on number of years worked as taxi drivers or forecourt attendants. A questionnaire was designed to investigate the health status of the participants. Blood samples were collected by medical professionals and analyzed for metal concentrations by ICP-MS.

**Results:** A positive correlation between number of years worked and Pb blood concentrations was found. The highest Pb concentration (60.2 μg/L) was observed in a forecourt attendant who had worked 11 to 20 years, and the average Pb concentration in this group (24.5 μg/L) was significantly higher (p<0.05) than in forecourt attendants who had worked 2 to 5 years (10.4 μg/L). Some individuals had elevated concentrations of manganese, arsenic, cadmium, chromium and cobalt, yet not significantly elevated at the group level. The blood levels of arsenic appeared to be related to smoking. Mood swings, dizziness, headaches and tiredness were reported by the participants.

**Conclusion:** Blood Pb concentrations in petrol station forecourt attendants and taxi drivers exposed to petrol are elevated and correlate to exposure time. A health monitoring program should be erected for all individuals working in these industries, and preventive measures should be implemented to eliminate metal exposure from petrol.

## 1. Introduction

Lead (Pb) has been added to gasoline since the 1920s to improve combustion engine performance. The use of leaded gasoline was officially banned in South Africa in 2006 (1). Before the ban, emissions from vehicles were considered the main source of human Pb exposure due to the use of tetraethyl lead gasoline (2), and such emissions were one of the major contributors of Pb and other toxic heavy metals into the environment (3-5). Tires, lubricating oil and gasoline (petrol) may also contribute to the atmospheric pollution (4). Reports have shown that vehicular emissions increase the concentration of heavy metals such as lead (Pb), arsenic (As), cadmium (Cd) and chromium (Cr) in soil and in the environment, and this has been attributed to petrol use in most cases (1). On the average between 1990 and 2013, a total of 250,000 deaths in Africa were linked to vehicular emissions (6) evoking the need for regulations.

Petroleum products pollute the air, water, soil, and vegetation and affect human health adversely. The toxic metals found in petrol products, such as lead, manganese, and nickel, are major public health concerns as their environmental concentrations continue to rise. Skin irritation, eye irritation, dizziness, headache, nausea, confusion and in extreme situations coma, cardiac arrythmia and death are all symptoms of petrol vapour exposure. Due to the widespread use of petroleum, unintentional acute discharges of metals from petroleum and its products can occur practically anywhere (7). The International Agency for Research on Cancer (IARC) reported in 1989 (8) that toxicity from petroleum-related products poses a health risk to humans. Many of the chemicals present in petroleum show cancerogenic properties (9).

The concentration of metals such as Pb, Mn and Cr in blood are often elevated in individuals occupationally exposed to petrol vapour. Blood concentrations of Pb in individuals directly exposed to gasoline while working as petrol station forecourt attendants (a.k.a. petrol station assistants) at different fuel stations in Basrah, Iraq, were significantly higher than in a control group mainly consisting of rural farmers (10). The average blood Pb level for the US population during the period when Pb was used in US gasoline was 3 to 5 times higher than the acceptable limit (11). High concentrations of Pb and Cd were found in occupationally exposed individuals working as motor mechanics, generator mechanics, and petrol station forecourt attendants in Nigeria when compared to a control group suggesting that such occupational exposure increases the blood level of Pb and Cd (12). Combustion of fossil fuels has been identified as one of the most significant sources of atmospheric Cd (12). Cadmium is also a by-product of Pb refining. Lead and Cd have a high absorption rate that ranges between 50 and 60% when inhaled from air. When ingested, the absorption rate is 3% for lead and 10% for cadmium (13).

The world demand for petrol has increased along with the growth of the global population and the expansion of the vehicle industry, leading to an increased occupational petrol exposure specifically in African countries (14-16). Petroleum-related toxicities pose a substantial health risk to humans (15, 16) and Pb accounts for 0.6% of the global burden of disease (17). Lead poisoning occurs through inhibition of haemoglobin synthesis, acute and chronic damage to the central and peripheral nervous system, cardiovascular and reproductive system affection and kidney dysfunction (18-20). When Pb ions bind to proteins they can induce misfolding and aggregation, leading to protein dysfunction and possible pathogenesis of protein aggregation diseases such as Alzheimer’s disease (21). In addition, psychosis can manifest from both acute and chronic Pb intoxication (22). Other petrol related metals such as As can cause symptoms such as nausea and vomiting, reduced nerve function and skin changes (23, 24). Manganese (Mn), which is now replacing Pb as an additive in some petrol brands to increase the octane number and to act as an anti-knock agent, also shows neurotoxic properties at high concentrations (25, 26).

Information on concentrations of toxic metals in the blood will provide information about human health status and the burden of pollutants in the environment and human health status (27). Despite the well-known toxicity of Mn and Pb and their extensive use as fuel additives, data on occupational metal exposure in Africa has not yet been fully explored, and if available details are scarce. The introduction of methylcyclopentadienyl manganesetricarbonyl (MMT) in petrol may also increase the levels of Mn in the environment. Manganese in physiological concentrations is essential for human health, unlike Pb, which is a non-essential metal without any function in biological systems (20). Mn and Pb vapors and fine particles are easily inhaled, thereby exposing forecourt attendants and other petrol related workers to these neurotoxic metals mainly via the respiratory exposure route, in addition to dermal and enteric exposures. Occupational exposure to metals is a growing public health concern in both developed and developing countries. The link between chemical exposure in the environment, body load, and potential harmful health effects is being extensively investigated using human biomonitoring techniques (28).

The aim of the current study was to determine concentrations of Pb, Mn and other metals in blood from forecourt attendants and taxi drivers before and after the introduction of unleaded petrol in South Africa in 2006, and to evaluate possible health effects of exposure to these metals. These cohorts were chosen because of their daily routines which involved close contact with petrol, most commonly through petrol dispersion.

## 2. Materials and methods

### 2.1 Participants

The study was conducted in Gauteng, a suburb in Pretoria, South Africa. Ethical approval was obtained from the Sefako Makgatho Health Sciences Research Committee (SMUREC/S/91/2020:PG). The study involved individuals who used petrol (taxi drivers) or sold petrol (petrol station forecourt attendants), either previously employed in these occupations or currently employed. As controls served individuals without any petrol contact on a daily basis, i.e. students or workers who reached their workplace on foot and who lived far away from any of the petrol stations studied. Within the study group (n=95) there were 38 forecourt attendants, 21 taxi drivers, and 36 controls. All participants signed a written consent form. The study groups were divided into subgroups based on their years of occupation, as shown in Tables 1-4.

**Table 1.**
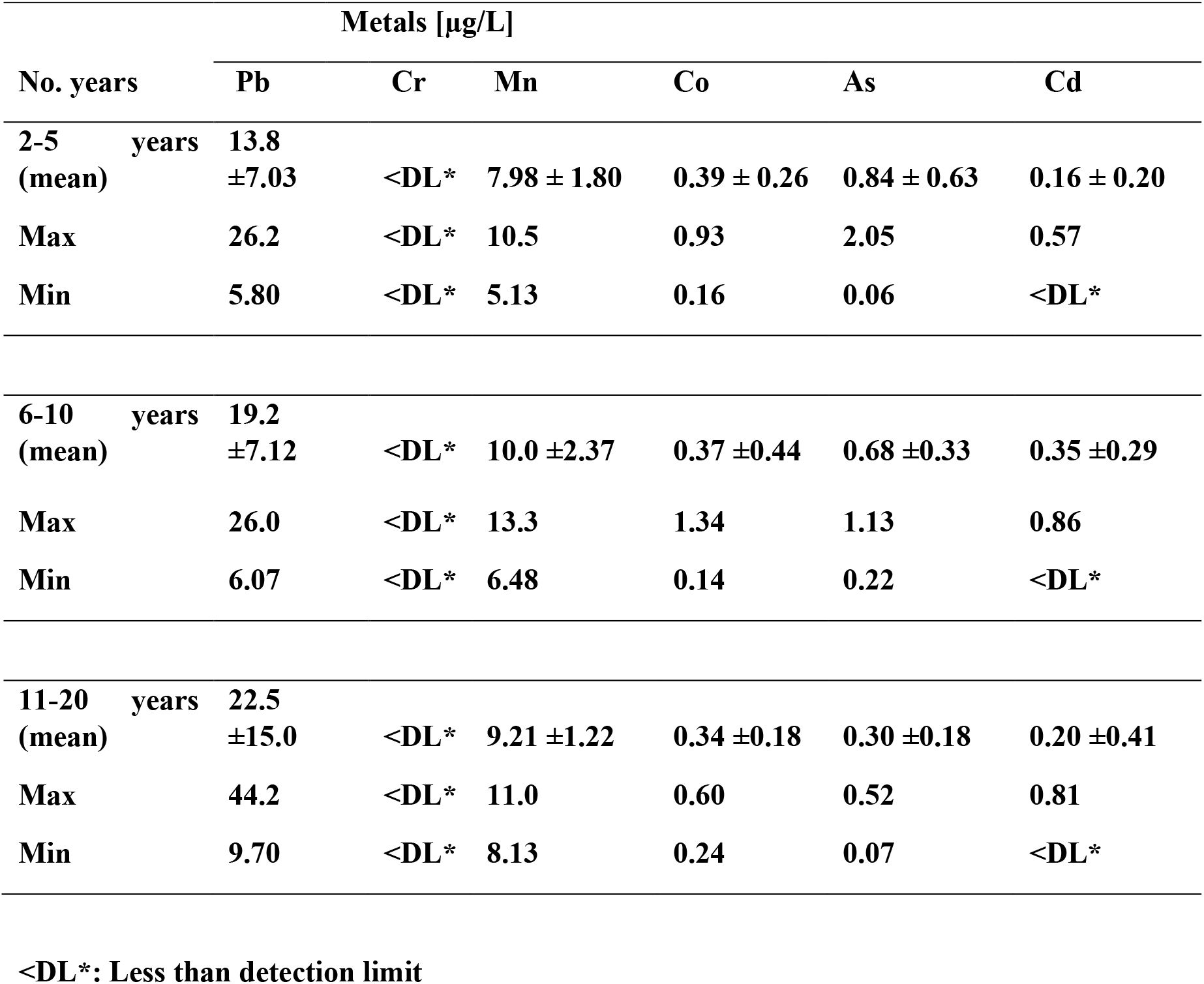
The mean and standard deviations of metal/metalloid concentrations (μg/L) in the blood of taxi drivers according to the number of years worked in the taxi industry.

**Table 2.** The mean and standard deviations of metal/metalloid concentrations (μg/L) in the blood of petrol station forecourt attendants according to the number of years worked in the industry.

| No. years | Metals [$\mu\text{g/L}$] | | | | | |
| --- | --- | --- | --- | --- | --- | --- |
|  | Pb | Cr | Mn | Co | As | Cd |
| **2-5 years** |  |  |  |  |  |  |
| **(mean)** | **10.4±5.39** | **0.07±0.27** | **9.71±2.71** | **0.69±0.71** | **0.93±0.68** | **0.10±0.16** |
| **Max** | **28.54** | **1.08** | **14.91** | **2.91** | **2.39** | **0.65** |
| **Min** | **5.21** | **<DL\*** | **5.32** | **0.08** | **<DL\*** | **<DL\*** |
| **6-10 years** |  |  |  |  |  |  |
| **(mean)** | **12.6±5.11** | **0.21±0.52** | **8.96±2.47** | **0.44±0.29** | **1.05±0.93** | **0.16±0.21** |
| **Max** | **23.8** | **1.28** | **14.0** | **1.09** | **3.55** | **0.63** |
| **Min** | **5.04** | **<DL\*** | **5.11** | **0.12** | **<DL\*** | **<DL\*** |
| **11-20 years** |  |  |  |  |  |  |
| **(mean)** | **24.5±18.2** | **0.29±0.74** | **9.16 ± 2.24** | **0.38** | **0.81±0.61** | **0.46±0.81** |
| **Max** | **60.2** | **2.11** | **12.8** | **0.69** | **1.62** | **0.93** |
| **Min** | **12.4** | **<DL\*** | **6.38** | **0.19** | **0.15** | **0.15** |
| **<DL\*: Less than detection limit** |  |  |  |  |  |  |

**Table 3.** The mean and standard deviations of metal/metalloid concentrations (μg/L) in the blood of the control group.

| | Metals [$\mu\text{g/L}$] | | | | | |
| --- | --- | --- | --- | --- | --- | --- |
|  | Pb | Cr | Mn | Co | As | Cd |
| **Mean ± SD** | **16.9 ± 9.2** | **0.5 ± 1.6** | **8.94 ± 1.55** | **0.3 ± 0.3** | **0.7 ± 0.5** | **0.3 ± 0.3** |
| **Max** | **41.3** | **7.4** | **16.3** | **1.20** | **2.94** | **1.3** |
| **Min** | **4.37** | **<DL\*** | **5.00** | **0.10** | **0.10** | **<DL\*** |

<DL\*: Less than detection limit

**Table 4.** Clinical symptoms reported by taxi drivers and petrol station forecourt attendants (n=59), presented according to number of years worked.

| Years worked | Symptoms |  |  |
| --- | --- | --- | --- |
|  | Headaches | Irritation/mood swings | Dizziness |
| **2-5** | 4 | 10 | 3 |
| **6-10** | 8 | 13 | 7 |
| **11-20** | 11 | 8 | 8 |
| **Total** | **n=23 (39 %)** | **n=31 (53 %)** | **n=18 (31 %)** |

Each participant received a structured questionnaire to fill out, with questions covering sociodemographic status, number of years working in their occupation, smoking habits, and health status. Before blood sampling, blood pressure was measured by a qualified health practitioner to ensure that the participants were neither hypertensive nor hypotensive. Each participant was also asked before sampling if they had any history of anemia. Blood was collected from the antecubital vein by a qualified health worker from Dr. George Mukhari Academic Hospital in Pretoria, using relevant and safe reference materials. Blood was collected in standard purple top EDTA vacutainer plastic tubes. Samples were stored on ice and stored in a refrigerator (minus 20 °C) awaiting further analysis. The blood sampling took place in a secluded and clean environment at each of the petrol stations and at the different motor parks. A private space was provided for the nurse and the participant away from their colleagues during work hours. The control group followed the same procedure albeit in the privacy of their homes.

#### 2.2 Metal concentration analysis

Analysis of the blood samples for the determination of metal concentrations took place at an accredited laboratory in South Africa. An Agilent 7700 series inductively coupled plasma– mass spectrometry (ICP–MS) instrument was used (29). Perchloric and nitric acid in a 1:3 ratio was used for the digestion procedure: 3 mL of perchloric acid and 9 mL of suprapure (Merck) concentrated nitric acid was added to 1 g of each sample in a quartz tube and vortexed for 30 s. Blood samples were digested with 0.5% HCl and 1.5% HN0_3_ in purified water. The extract was then diluted, made up to volume, centrifuged, and the supernatants decanted for metal concentrations reading using ICP-MS.

For the purpose of quality assurance and reliability of the results, samples were analyzed in triplicate and both internal quality control standards and certified reference materials - ClinChek® - specific to each metal were used to verify and validate the results. Samples were spiked with metals in known concentrations for reference, and reagent blanks were run with every batch of the samples analyzed and results monitored as per the ISO 17025 guidelines. For all six metals measured, i.e. As, Cd, Co, Cr, Mn and Pb, the Limit of Detection (LOD) was 0.2 μg/L, and the Limit of Quantification (LOQ) was 0.6 μg/L.

### 2.3 Statistical analysis

The SPSS 28.0 statistical software was used for statistical analysis. One-way ANOVA was performed to see if the differences obtained in the metal concentrations from each group were significant or not. When values obtained were lower than the detection limit, they were divided by 2 (LOD/2). The statistical significance level was set to p<0.05.

## 3. Results and discussion

The participants in this study were predominantly male as female petrol station forecourt attendants and taxi drivers are rare in Pretoria. Other studies in Africa have noted a similar skew between males and females (30, 31) where only two females (1.1%) participated. The studies confirmed that these jobs are demanding physically due to the equipment used and therefore viewed as jobs for men (30, 31). The blood metal concentrations of the different groups are presented in Tables 1, 2 and 3.

### 3.1 Lead: blood concentrations and health aspects

Blood Pb concentrations in individual forecourt attendants and taxi drivers ranged from 5.04 μg/L to 60.2 μg/L (Tables 1 and 2). Although no occupation group showed Pb concentrations significantly different from that in the control group (16.9 μg/L; Table 3), there is a clear trend that both taxi drivers and forecourt attendants display higher Pb concentrations with more years worked (Tables 1 and 2). Thus, the highest average Pb concentration (24.5 μg/L; Table 2) was found in petrol station forecourt attendants who had worked between 11 and 20 years. These people had also been smoking for many years. The measured Pb concentration was significantly higher (p<0.05) than the average Pb concentration in forecourt attendants who had worked between 2 and 5 years, i.e. 10.4 μg/L (Table 2). This dose-response relationship indicates that the elevated Pb concentrations found are related to occupational exposure to Pb.

A similar trend like the one reported in our study was previously found in roadside technicians in Lagos, Nigeria, where it was reported that blood Pb concentrations were higher in the technicians who had spent more than 10 years in their job, when compared to those who had spent 2 - 5 years in their job (30). From the results by Saliu et al. (30) and our results, it is evident that an increase in blood Pb concentration is correlated to the number of years of Pb exposure in these jobs. It should be recalled that the introduction of unleaded fuel started in South Africa in 2005. The Pb concentrations reported in our study are lower than those reported from Lagos, Nigeria (30) where the blood Pb levels of the roadside mechanics exposed to leaded fuel were much higher than the WHO recommended limit, i.e., 50 μg/L (32). Lead in blood may reside long term in equilibrium with bone Pb and adverse health effects can occur many years after Pb exposure (11). The McFarland study pointed out that although the participants in their study were exposed when they were young profound health effects may precipitate later in life. Animal and epidemiological studies have shown that early exposure may affect multiple organs ultimately leading to impaired cognitive ability and emotional regulation (11).

Irritation and emotional disturbances were reported by 53% of the current participants with high blood Pb concentrations (Table 4). Around one third, i.e. 39% and 32%, also complained about frequent headache and dizziness, respectively (Table 4). Health issues associated with Pb exposure and reports from previous studies have shown that irritation, stomach ache, diarrhoea, colic, distractibility, and lethargy are among the mild symptoms of Pb poisoning, while severe Pb exposure may affect haematopoiesis and the nervous, cardiovascular, urinary, and reproductive systems (20, 33). The only symptom mentioned by most participants in our study was mood swings in those with high blood Pb (Table 4).

### 3.2 Manganese: blood concentrations and health aspects

The Mn concentrations in individual forecourt attendants and taxi drivers ranged from 5.11 μg/L to 14.9 μg/L in our study (Tables 1 and 2). No occupation group had average Mn concentrations that were significantly different from that of the control group (8.9 μg/L; Table 3). The highest Mn concentration was recorded in the blood of a taxi driver who had worked for only two years. The differences in Mn concentration between subgroups of petrol station forecourt attendants and taxi drivers were not significant. A previous study has reported blood Mn concentrations of 14.2 to 22.6 μg/L in occupationally exposed individuals in Nigeria (34), a range slightly higher than the values reported in our study.

The results of our study are also similar to those reported by Mehrifar et al. (35) on welders exposed to Mn, where Mn concentrations of 18.3 ± 5.84 μg/L were found. Neurobehavioral symptoms were also significantly higher in welders compared to controls in the Mehrifar study (35), which concluded that welders are more prone to Mn toxicity by elevated Mn concentrations in their blood due to the nature of their work, as chemicals high in Mn are easy to inhale. Manganese creates harmful oxygen radicals (36), induces protein aggregation and misfolding (37) and accumulates in the brain where it may cause central nervous system disorders and psychiatric disorders (26, 38). The addition of Mn in the form of MMT to petrol has been reported as a harmless additive producing Mn exposures comparable to the normal Mn background and may not create health problems (39). However, Mn exposure from other sources like drinking water or food apart from Mn exposure from MMT petrol may potentially increase the levels of Mn in an individual adding up to toxic levels. A limited increase in Mn exposure was observed in taxi drivers from Canada (25). According to the findings by Zayed et al. (25), the control group’s concentration was lower than that of the study group and below the WHO limit. The low levels of Mn reported from the control group used in the present study supported the initial observation reported by Zayed et al. (25).

### 3.3 Arsenic: blood concentrations and health aspects

The metalloid arsenic was not detected in most of the participants, and no occupation group (Tables 1 and 2) had average As concentrations that were significantly different from that of the control group (i.e. 0.7 μg/L; Table 3). The highest concentration of As, 3.55 μg/L, was recorded in the blood from a forecourt attendant who had been working for more than 7 years. The overall As concentrations did not correlate to number of years worked, and some data was missing as As was not measured in all individuals. However, As had positive correlation with smoking (data not shown), and we therefore argue that the main reason for the traceable levels of As in the studied individuals was from the involvement with smoking. WHO reported in 2002 (40) that As is on the list of ten chemicals of major public health concern and it has been noted that tobacco plants can absorb inorganic arsenic directly from the soil (21). Cigarette smoking has also been linked to an increase in blood As levels, which may increase the risk of heart disease, skin lesions, bladder cancer, and lung cancer (41).

### 3.4 Cadmium: blood concentrations and health aspects

Cadmium was not detected in most of the participants in the study groups or in the control group (Tables 1-3). No significant differences in Cd concentrations were found between subgroups, or when compared to the mean Cd concentration in the control group (0.3 μg/L: Table 3). Cadmium concentrations measured in some individuals were high (Tables 1-3), but still comparable to Cd concentrations found in other studies where individuals were occupationally exposed to Cd (Liao et al. 2019). The concentrations of Cd from the participants did not follow any specific pattern. The highest Cd concentrations were found in (a) a forecourt attendant with a blood Cd concentration of 0.93 μg/L, and (b) a taxi driver with a blood Cd concentration of 0.86 μg/L who had been working for more than 7 years. These concentrations are lower than those reported in a study of people living near a zinc smelter (42). Human exposure to Cd occurs mainly from consumption of Cd contaminated food, active and passive inhalation of tobacco smoke, and inhalation by industrial workers in a range of industries as well as inhalation of vehicular Cd emissions. Cadmium is known to promote nephrotoxicity and prolonged exposure to Cd can lead to its accumulation in the kidney and may cause neotubular dysfunction (20, 43). The toxic effects of Cd on the central nervous system, especially in conjunction with other neurotoxic metals such as Pb, have been extensively reviewed (42, 44). Some promising results have been achieved with the disaccharide trehalose in alleviating Cd induced brain damage in rats (43, 45).

### 3.5 Chromium: blood concentrations and health aspects

Chromium was not detected in the blood from any of the taxi drivers (Table 1), and only in some of the forecourt attendants (Table 2) and some controls (Table 3). The detected Cr concentrations were generally below 1 μg/L, although there were individuals with Cr concentrations as high as 2.11 μg/L among the forecourt attendants (Table 2), and 7.4 μg/L in the control group (Table 3). Because of these two individuals with unusually high Cr concentrations, and the overall low number of individuals with detectable amounts of Cr, the statistics for the Cr concentrations are somewhat unreliable, and no significant differences in blood Cr concentrations were observed between any of the groups. Chromium may be released into the environment from the exhaust of diesel engines, products of friction braking systems, and as a result of asphalt road wear (46). Generally, the Cr concentrations reported in this study (Tables 1-3) were lower than those reported from Nigeria, where values ranged from 12.75 to 16.44 μg/L (34).

### 3.6 Cobalt: blood concentrations and health aspects

Cobalt concentrations ranged from 0.80 μg/L to 2.91 μg/L in the petrol station forecourt attendants (Table 2), 0.14 μg/L to 1.34 μg/L in the taxi drivers (Table 1), and 0.10 μg/L to 1.20 μg/L in the control group (Table 3). Both Cr and Co exposure may cause lung cancer, and Co has a well-known function as a cofactor for vitamin B12 (20).

### 3.7 Strengths and weaknesses of the study

This study provides a unique set of data from a region of the world where occupational exposure to Pb and other neurotoxic metals is high. The choice of petrol station forecourt attendants and taxi drivers for study provides insights into respiratory exposure routes for metals from handling of petroleum. Smoking habits of some individuals might influence the levels of metals in the blood (21). The study did not examine in detail the adverse effect of all pollutants affecting the individuals who participated in this study, however the analysis of the questionnaires unveiled complaints about headache, dizziness and tiredness. The weaknesses of a questionnaire approach in obtaining these results are acknowledged.

## 4. Conclusions

Blood concentrations of lead (Pb) are elevated in taxi drivers and petrol station forecourt attendants in Pretoria, South Africa. The blood Pb concentrations increase with employment time, indicating a dose-response effect related to occupational exposure to petrol. Arsenic, cadmium, chromium, cobalt, and manganese concentrations are elevated in some individuals, but not significantly elevated at the group level. Preventive measures to eliminate metal exposure from petrol are necessary, together with frequent medical examinations and monitoring of the health of exposed individuals.

## Data Availability

All data produced in the present work are contained in the manuscript

## Acknowledgments

Support to SW from the Magnus Bergvall foundation and support to PMR from the Kamprad Research Foundation, the Magnus Bergvall Research Foundation, the Ulla-Carin Lindquist Foundation for ALS Research and the Karolinska Institutet IMM strategic grants is gratefully acknowledged. The support from NRF, South Africa is also greatly appreciated.

## References

1. Olowoyo JO, Lion N, Unathi T, Oladeji OM. Concentrations of Pb and Other Associated Elements in Soil Dust 15 Years after the Introduction of Unleaded Fuel and the Human Health Implications in Pretoria, South Africa. Int J Environ Res Public Health. 2022;19(16).

2. Kayee J, Bureekul S, Sompongchaiyakul P, Wang XF, Das R. Sources of atmospheric lead (Pb) after quarter century of phasing out of leaded gasoline in Bangkok, Thailand. Atmos Environ. 2021;253.

3. Dignam T, Kaufmann RB, LeStourgeon L, Brown MJ. Control of Lead Sources in the United States, 1970-2017: Public Health Progress and Current Challenges to Eliminating Lead Exposure. J Public Health Manag Pract. 2019;25 Suppl 1, Lead Poisoning Prevention(Suppl 1 LEAD POISONING PREVENTION):S13–S22.

4. Nawrot N, Wojciechowska E, Rezania S, Walkusz-Miotk J, Pazdro K. The effects of urban vehicle traffic on heavy metal contamination in road sweeping waste and bottom sediments of retention tanks. Sci Total Environ. 2020;749:141511.

5. Levin R, Zilli Vieira CL, Rosenbaum MH, Bischoff K, Mordarski DC, Brown MJ. The urban lead (Pb) burden in humans, animals and the natural environment. Environ Res. 2021;193:110377.

6. Raje F, Tight M, Pope FD. Traffic pollution: A search for solutions for a city like Nairobi. Cities. 2018;82:100–7.

7. Anderson AR, Centers for Disease C. Health Effects of Cut Gas Lines and Other Petroleum Product Release Incidents - Seven States, 2010-2012. MMWR Morb Mortal Wkly Rep. 2015;64(22):601–5.

8. IARC. Occupational Exposures in Petroleum Refining; Crude Oil and Major Petroleum Fuels. 1989.

9. Onyije FM, Hosseini B, Togawa K, Schuz J, Olsson A. Cancer Incidence and Mortality among Petroleum Industry Workers and Residents Living in Oil Producing Communities: A Systematic Review and Meta-Analysis. Int J Environ Res Public Health. 2021;18(8).

10. Al-Rudainy LA. Blood lead level among fuel station workers. Oman Med J. 2010;25(3):208–11.

11. McFarland MJ, Hauer ME, Reuben A. Half of US population exposed to adverse lead levels in early childhood. Proc Natl Acad Sci U S A. 2022;119(11):e2118631119.

12. Elehinafe FB, Mamudu AO, Okedere OB, Ibitioye A. Risk assessment of chromium and cadmium emissions from the consumption of premium motor spirit (PMS) and automotive gas oil (AGO) in Nigeria. Heliyon. 2020;6(11):e05301.

13. Faroon O, Ashizawa A, Wright S, Tucker P, Jenkins K, Ingerman L, et al. Toxicological Profile for Cadmium. Atlanta (GA)2012.

14. Campo L, Rossella F, Mercadante R, Fustinoni S. Exposure to BTEX and Ethers in Petrol Station Attendants and Proposal of Biological Exposure Equivalents for Urinary Benzene and MTBE. Ann Occup Hyg. 2016;60(3):318–33.

15. Kasemy ZA, Kamel GM, Abdel-Rasoul GM, Ismail AA. Environmental and Health Effects of Benzene Exposure among Egyptian Taxi Drivers. J Environ Public Health. 2019;2019:7078024.

16. Qafisheh N, Mohamed OH, Elhassan A, Ibrahim A, Hamdan M. Effects of the occupational exposure on health status among petroleum station workers, Khartoum State, Sudan. Toxicol Rep. 2021;8:171–6.

17. Liao KW, Pan WH, Liou SH, Sun CW, Huang PC, Wang SL. Levels and temporal variations of urinary lead, cadmium, cobalt, and copper exposure in the general population of Taiwan. Environ Sci Pollut Res Int. 2019;26(6):6048–64.

18. Wani AL, Ara A, Usmani JA. Lead toxicity: a review. Interdiscip Toxicol. 2015;8(2):55–64.

19. Assi MA, Hezmee MN, Haron AW, Sabri MY, Rajion MA. The detrimental effects of lead on human and animal health. Vet World. 2016;9(6):660–71.

20. Nordberg G, Costa M. Handbook on the toxicology of metals, 5th ed. London, U.K.: Academic Press; 2021.

21. Wallin C, Sholts SB, Österlund N, Luo J, Jarvet J, Roos PM, et al. Alzheimer’s disease and cigarette smoke components: effects of nicotine, PAHs, and Cd(II), Cr(III), Pb(II), Pb(IV) ions on amyloid-beta peptide aggregation. Sci Rep. 2017;7(1):14423.

22. Duruibe JO, Ogwuegbu MOC, Egwurugwu JN. Heavy metal pollution and human biotoxic effects. Int J Phys Sci. 2007;2(5):112–8.

23. Smith AH, Lingas EO, Rahman M. Contamination of drinking-water by arsenic in Bangladesh: a public health emergency. Bull World Health Organ. 2000;78(9):1093–103.

24. Jaishankar M, Tseten T, Anbalagan N, Mathew BB, Beeregowda KN. Toxicity, mechanism and health effects of some heavy metals. Interdiscip Toxicol. 2014;7(2):60–72.

25. Zayed J, Mikhail M, Loranger S, Kennedy G, L’Esperance G. Exposure of taxi drivers and office workers to total and respirable manganese in an urban environment. Am Ind Hyg Assoc J. 1996;57(4):376–80.

26. Aschner M, Guilarte TR, Schneider JS, Zheng W. Manganese: recent advances in understanding its transport and neurotoxicity. Toxicol Appl Pharmacol. 2007;221(2):131–47.

27. Yedomon B, Menudier A, Etangs FLD, Anani L, Fayomi B, Druet-Cabanac M, et al. Biomonitoring of 29 trace elements in whole blood from inhabitants of Cotonou (Benin) by ICP-MS. J Trace Elem Med Biol. 2017;43:38–45.

28. Cerna M, Krskova A, Cejchanova M, Spevackova V. Human biomonitoring in the Czech Republic: an overview. Int J Hyg Environ Health. 2012;215(2):109–19.

29. Olowoyo JO, Macheka LR, Mametja PM. Health Risk Assessments of Selected Trace Elements and Factors Associated with Their Levels in Human Breast Milk from Pretoria, South Africa. Int J Environ Res Public Health. 2021;18(18).

30. Saliu A, Adebayo O, Kofoworola O, Babatunde O, Ismail A. Comparative assessment of blood lead levels of automobile technicians in organised and roadside garages in Lagos, Nigeria. J Environ Public Health. 2015;2015:976563.

31. Aiyenigba BO. Effect of health education on the knowledge and safety practices of auto mechanics in Lagos state 2005.

32. WHO. WHO guideline for clinical management of exposure to lead. Geneva: World Health Organization; 2021.

33. des Planches L. Traité des maladies de plomb ou saturnines. Paris: Ferra; 1839.

34. Etsuyankpa MB, Ahmad AS, Mustapha S, Baba NM, Ndamitso MM, Elabor R. Levels of selected heavy metals in blood and urine of workers of Alkali Kongo Plaza GSM Market within Lafia Metropolis, Nasarawa State, Nigeria. Environ Challenges. 2022;9:100649.

35. Mehrifar Y, Bahrami M, Sidabadi E, Pirami H. The effects of occupational exposure to manganese fume on neurobehavioral and neurocognitive functions: An analytical cross-sectional study among welders. EXCLI J. 2020;19:372–86.

36. Roos E, Wärmländer SKTS, Meyer J, Sholts SB, Jarvet J, Gräslund A, et al. Amyotrophic Lateral Sclerosis After Exposure to Manganese from Traditional Medicine Procedures in Kenya. Biol Trace Elem Res. 2021;199(10):3618–24.

37. Wallin C, Kulkarni YS, Abelein A, Jarvet J, Liao Q, Strodel B, et al. Characterization of Mn(II) ion binding to the amyloid-beta peptide in Alzheimer’s disease. J Trace Elem Med Biol. 2016;38:183–93.

38. Antonini JM, Santamaria AB, Jenkins NT, Albini E, Lucchini R. Fate of manganese associated with the inhalation of welding fumes: potential neurological effects. Neurotoxicology. 2006;27(3):304–10.

39. Cooper WC. The health implications of increased manganese in the environment resulting from the combustion of fuel additives: a review of the literature. J Toxicol Environ Health. 1984;14(1):23–46.

40. Exposure To Arsenic: A Major Public Health Concern. Geneva: World Health Organization 2002. Report No.: WHO/CED/PHE/EPE/19.4.1.

41. Chen CL, Hsu LI, Chiou HY, Hsueh YM, Chen SY, Wu MM, et al. Ingested arsenic, cigarette smoking, and lung cancer risk: a follow-up study in arseniasis-endemic areas in Taiwan. JAMA. 2004;292(24):2984–90.

42. Jo H, Kim G, Chang J, Lee K, Lee C, Lee B. Chronic Exposure to Lead and Cadmium in Residents Living Near a Zinc Smelter. Int J Environ Res Public Health. 2021;18(4).

43. WHO. Preventing disease through healthy environments: exposure to cadmium: a major public health concern. Geneva: World Health Organization; 2019. Report No.: WHO/CED/PHE/EPE/19.4. 3.

44. Andrade VM, Aschner M, Marreilha Dos Santos AP. Neurotoxicity of Metal Mixtures. Adv Neurobiol. 2017;18:227–65.

45. Tang KK, Liu XY, Wang ZY, Qu KC, Fan RF. Trehalose alleviates cadmium-induced brain damage by ameliorating oxidative stress, autophagy inhibition, and apoptosis. Metallomics. 2019;11(12):2043–51.

46. Gromadzinska J, Wasowicz W. Health risk in road transport workers. Part I. Occupational exposure to chemicals, biomarkers of effect. Int J Occup Med Environ Health. 2019;32(3):267–80.

